# The Influence of Covid-19 Vaccine on Daily Cases, Hospitalization, and Death Rate in Tennessee: A Case Study in the United States

**DOI:** 10.1101/2021.03.16.21253767

**Authors:** Ali Roghani

## Abstract

The COVID-19 outbreak highlights the vulnerability to novel infections, and vaccination remains a foreseeable method to return to normal life. However, infrastructure is inadequate for the whole population to be vaccinated immediately. Therefore, policies have adopted a strategy to vaccinate the elderly and vulnerable population while delaying others. This study uses the Tennessee official statistic from the onset of COVID vaccination (17th of December 2021) to understand how age-specific vaccination strategies reduce daily cases, hospitalization, and death rate. The result shows that vaccination strategy can significantly influence the numbers of patients with COVID-19 in all age groups and lower hospitalization and death rates just in older age groups. The Elderly had a 95% lower death rate from December to March; however, and no change in the death rate in other age groups. The Hospitalization rate was reduced by 80% in this study cohort for people aged 80 or older, while people who were between 50 to 70 had almost the same hospitalization rate. The study indicates that vaccination targeting older age groups is the optimal way to avoid higher transmissions and reduce hospitalization and death rate for older groups.

## Introduction

COVID-19 Vaccination rollout in Tennessee started on December 17, 2020, and by March 3, 13.3% of the population had already received mRNA vaccines like the Pfizer BNT162b2 (Tozinameran) and the Moderna mRNA-1273(1,2). In addition to reduced interpersonal contact and physical distancing, vaccination programs positively influence controlling the virus’s spread and reducing deaths (3). While COVID cases and deaths had the highest rates in January 2021 in the U.S., manufacturers currently cannot cover the enormous demand. As the vaccine supply is limited, it is crucial to prioritize who gets the vaccine; therefore, groups at the highest risk of getting the virus or individuals who are seriously ill receive the vaccine first. Besides, Tennesseans are eligible for vaccines based solely on their age, and these age-based phases have run simultaneously with those at high-risk health conditions. This paper has modeled how the vaccination program in Tennessee is likely to impact COVID-19-related daily cases, deaths, and hospitalization among adults.

### Data

This study used publicly available data of COVID-19, including vaccination rates, positive cases, hospitalizations, and death from the health department of Tennessee. All data and code used in this study are available at https://www.tn.gov/health/cedep/ncov/data. This study used the data from the first date of vaccinations, December 17, 2020, to March 3, 2021. The rates were adjusted by data from U.S. Census Bureau (2019) (4). The data were stratified from 21 years, ten years interval, and the last age group was 81 and more.

## Result

During the first 78 days of vaccination in Tennessee, there were 953568 individuals vaccinated by the 1st dose. By that date, 495032 individuals received their 2nd dose of the vaccine. 18.2% and 30.3% of vaccines were for people of age 81+ and 71-80, respectively, which shows that half of the vaccination has been for those older than 71.

Figure 1 indicates Tennessean’s total percentage of those who received the first vaccination from December 17, 2020, to March 3, 2021. Less than 70 had a higher vaccination rate before January than age groups of 71-80 and 80+; however, from January to March 3, 40%, 36% of 80+, and 71-80 have been vaccinated receptively, and it shows most of the daily vaccines were used for these two age groups. Figure 2 shows that just 25 % of Tennesseans older than 81 received the second dosage of vaccine, and a small percentage of other age groups have been vaccinated. Although the age group of 71-80 had a very close rate of receiving the first dosage of vaccine with 81+ age groups, they received the second dosage similar to other age groups and considerably lower than their counterparts who are older than 81.

**Figure 1.**
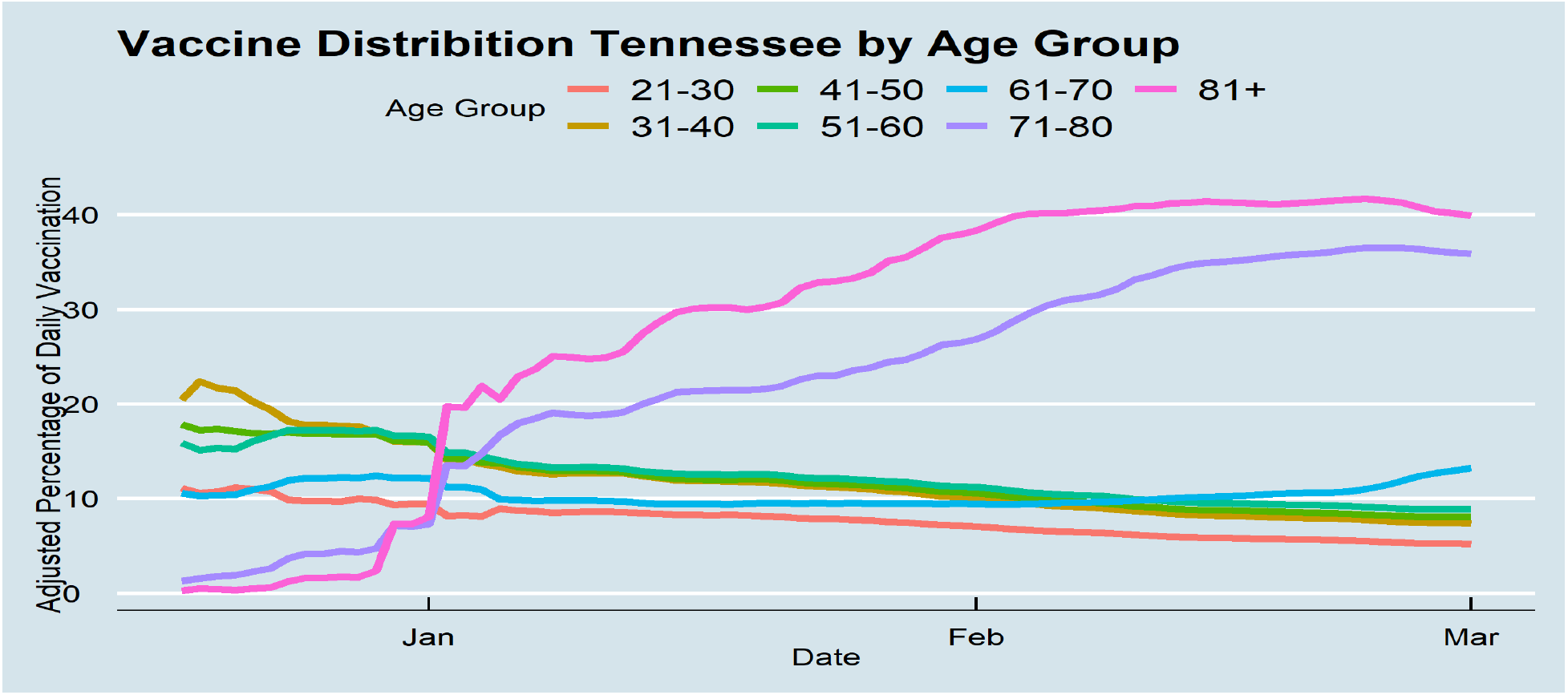

**Figure 2.**
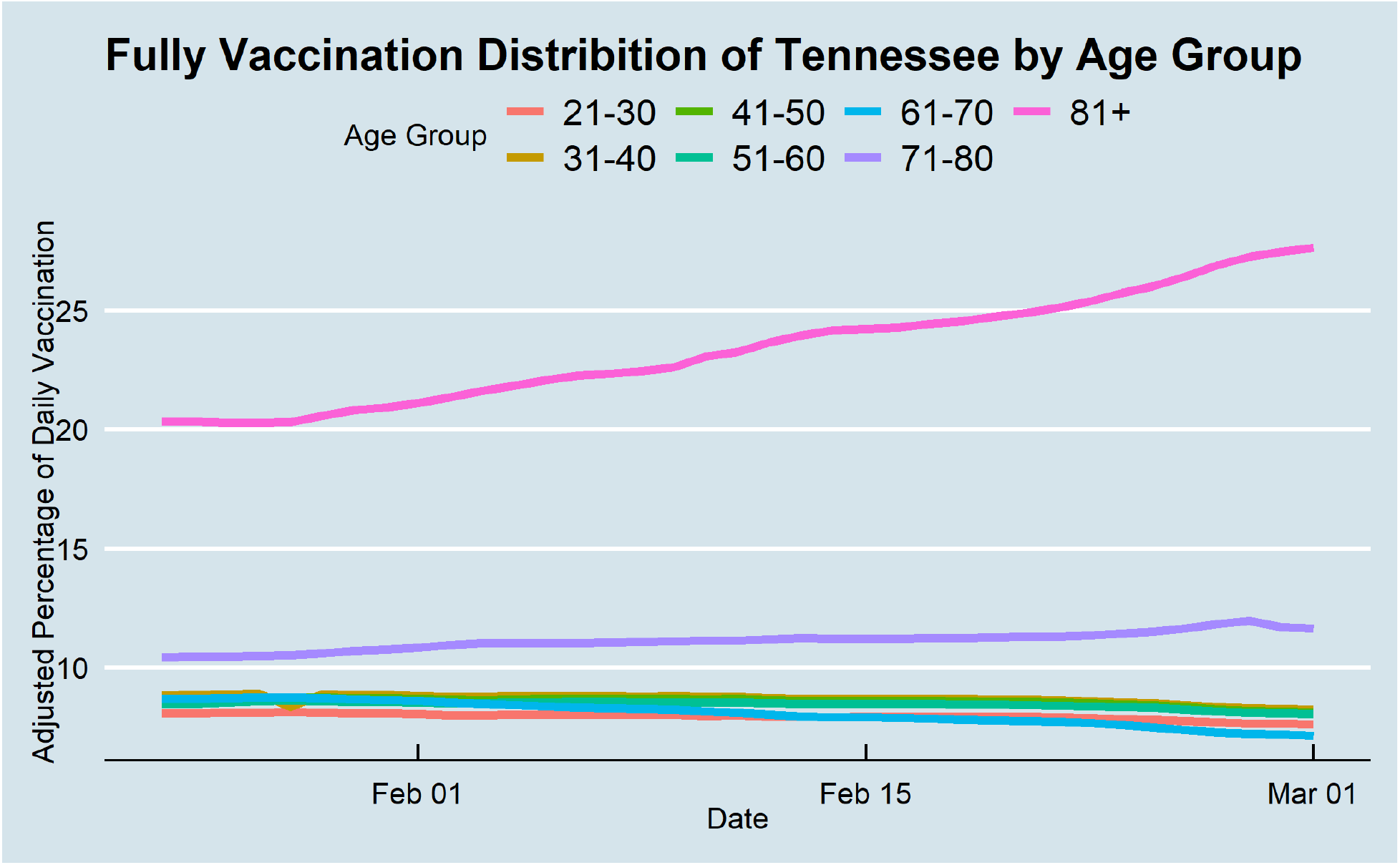

Daily cases of all age groups from the onset of vaccination to day 78 decreased inevitably (Figure 3). Daily cases were decreased from around 0.2% of the younger age group’s total population at the end of January to less than 0.05% at the end of the study period. This percentage was considerably higher for the older people in the study periods (from 0.1% to close to 0.01% daily cases). Before starting the vaccinations, older age groups were considerably had the highest hospitalizations rates. In contrast, their hospitalizations decreased at the end of the study period largely from 0.010% of Tennessee’s older population to 0.003%. There was no substantial change in other age groups’ hospitalization rates, although, on some days, age groups of 51-60 and 61-70 had high hospitalization rates. From the middle of February, age groups of 70+ did not experience high rates of hospitalization, and age group pf 51-60 had almost the highest rate of hospitalization. Lastly, vaccination decreased the death rates of 71+ age groups, while there was no change in death rates of other age groups during the study period. Although Tennesseans over 71 faced 0.015% of daily death at the end of 2020 by COVID, this percentage decreased substantially to 0.003% at the end of the study period. The results show that the gap between the older and younger was high before starting vaccination, but after vaccination the differences diminished, and all age groups had the same death rates.

**Figure 3.**
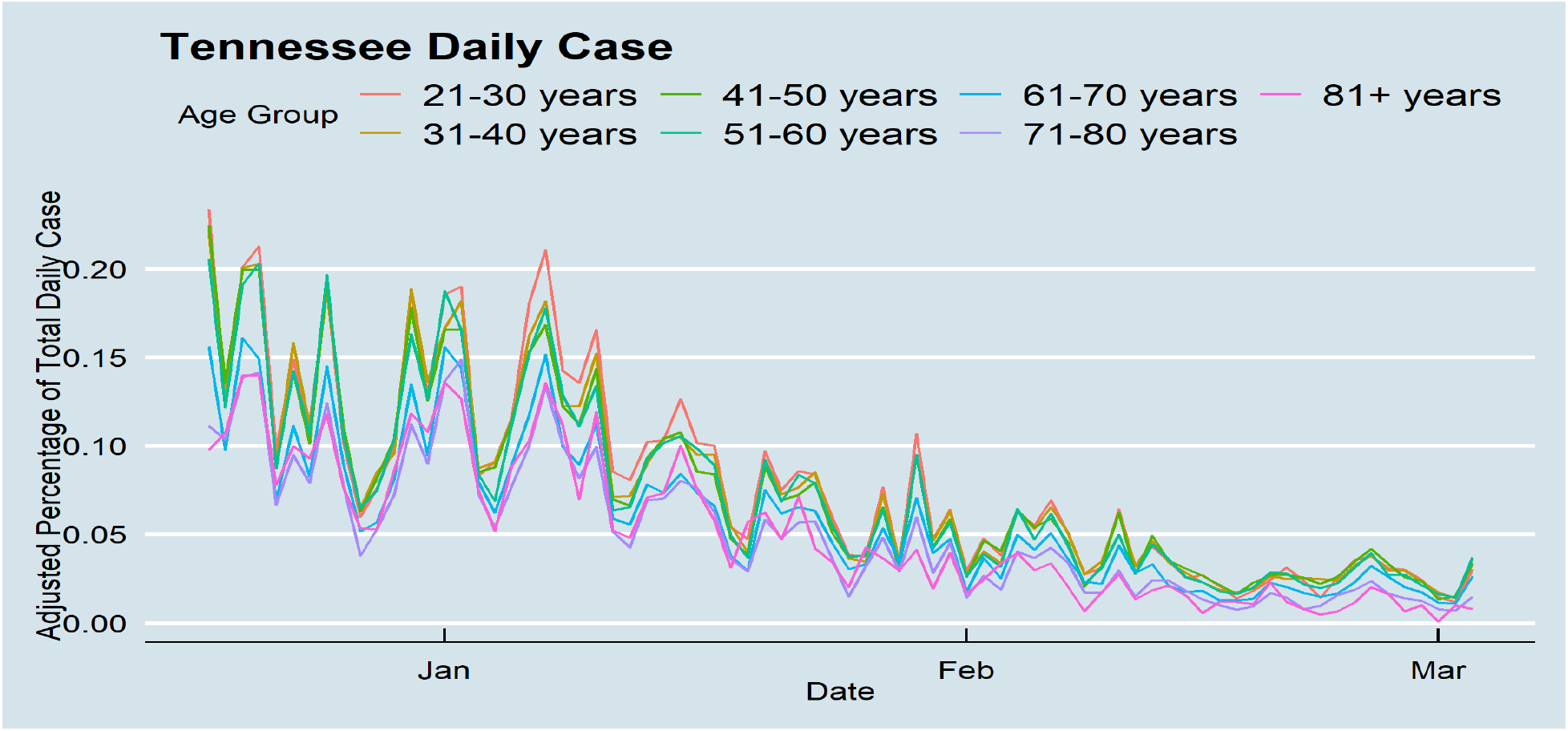

**Figure 4.**
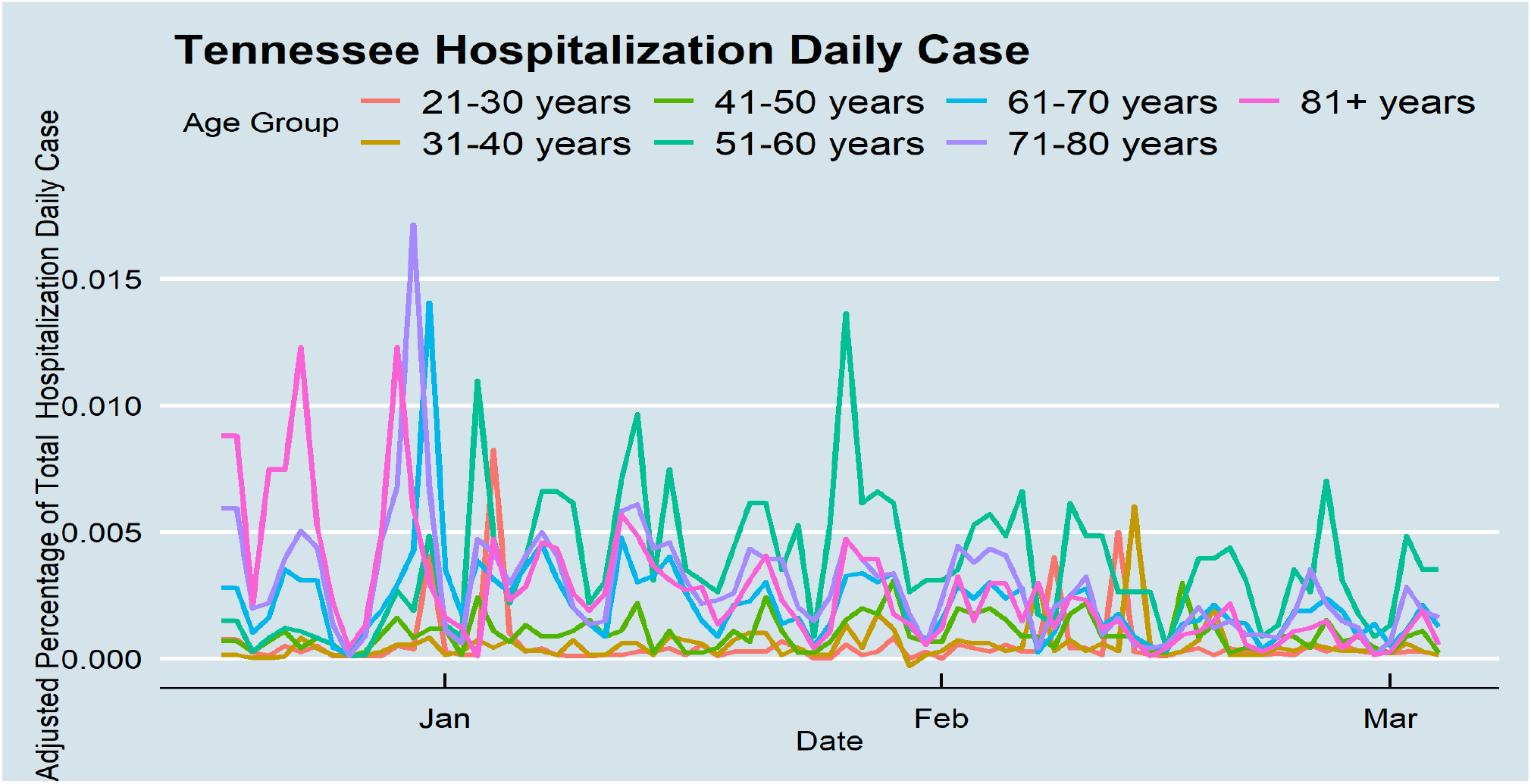

**Figure 5.**
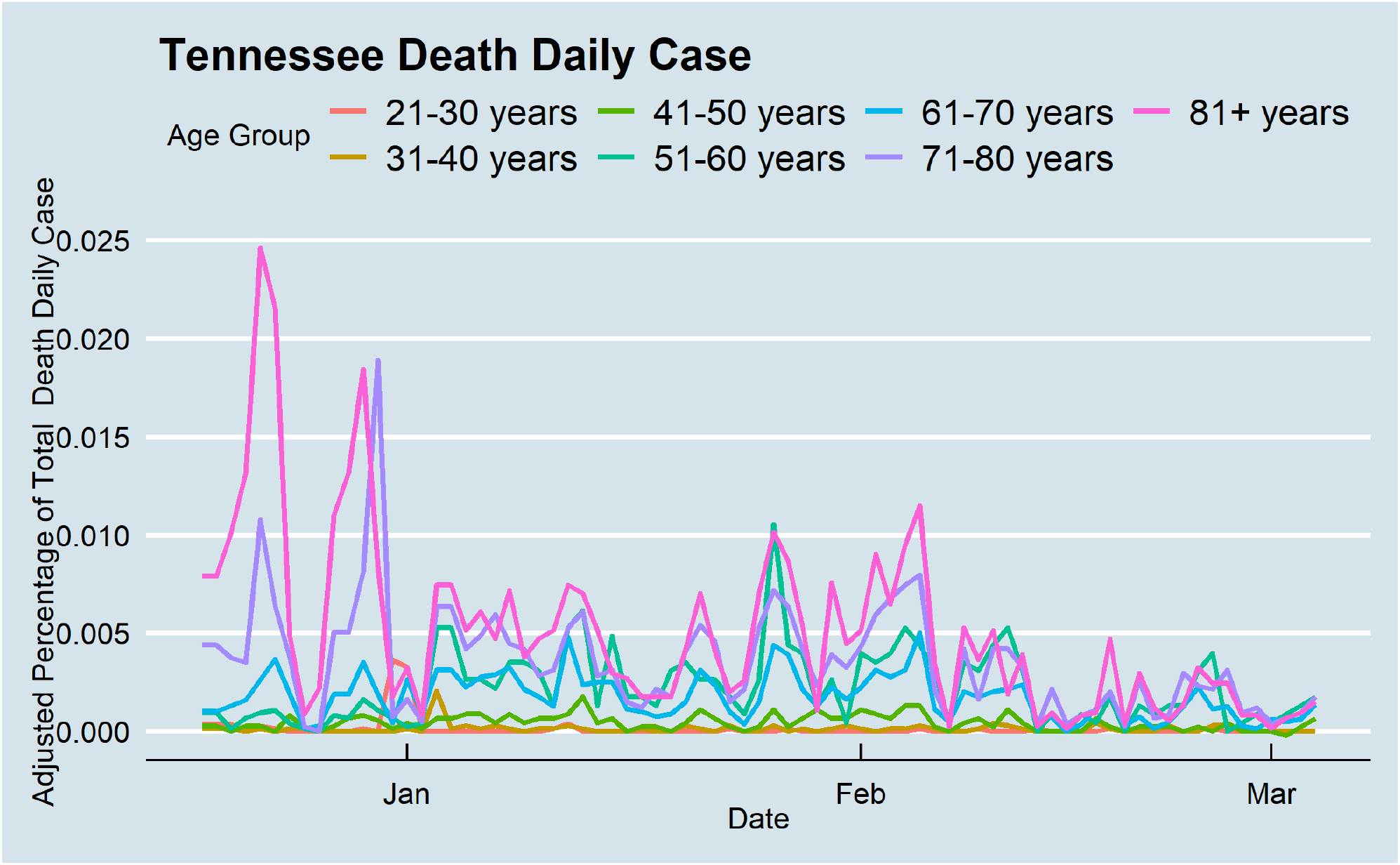

## Discussion

The COVID-19 is still spreading in the United States, and the hospitalization and death rate are high. Vaccines offer great hope for better conditions, but an effective vaccination strategy is needed to stop the pandemic and restore people’s everyday lives. Unfortunately, vaccine doses are being delivered slowly and sporadically, which means difficult for most people to be vaccinated right now, even if they are eligible. Based on the current policy, the high-risk groups such as first responders, the elderly, and individuals with high-risk health conditions should receive the vaccines first (5). In this study, I used the Tennessee official statistic from the onset of COVID vaccination (17th of December 2021) to understand how age-specific vaccination strategies change daily cases, hospitalization, and death rate. The mapping indicates that phase 1 of the vaccination strategy has the potential impacts to the numbers of patients in all age groups, lower hospitalization and death rate for elderly. The result demonstrates that more than half of vaccines were for people who are more than 70 years old, and it was an effective approach in blocking the transmission not only in the elderly population but also in other age groups. COVID-19 daily cases of older groups decreased to 90 % from the end of 2020 by the end of February 2021. Also, less than half of vaccines were used for less than 70, and they had less than 80% lower daily cases at the end of the study period. Although this study cannot confirm the association between the onset of vaccination and the considerable reduction in Covid-19 transmission in younger age groups, the statistics indicate a significant decrease in daily cases among Tennessean in all age groups. Moreover, 25% of people who were older than 81 received the vaccines, and around 10% of other age groups received the second dosage. However, this age group did not have better results than their counterparts in the 71-80 age group in hospitalizations and death rate. This study includes 78 days of vaccination statistics; thus, it is too early to conclude the second dosage’s influence. Future studies should consist of a longer period to have more precious results concerning the second dosage.

Vaccines lead to great hospitality and mortality reduction for older age groups in Tennessee. People who older than 80 had a 95% less death rate than in the middle of December. 71-80 age group death rate decreased during the study period; however, the 61-70 age groups had almost the same death rate from the middle of December to the end of February. The statistics show that there was no change in the death rate in other age groups. More than 80 years of Tennessean hospitalization reduced to 80 % in the study period, while people who were between 50 to 70 had almost the same hospitalization. It is essential to know individuals who were between 51-60 had the highest hospitalization rates in Tennessee. Although the data cannot identify people with higher risk, the higher hospitalization rate among the younger population implies health system in Tennessee could not identify people at higher risk efficiently. A previous study shows that a significant proportion of the people who had two or more chronic conditions simultaneously are more likely to be hospitalized by SARS-CoV-2 (9).

The findings should be considered in the context of several data limitations. I did not derive individual-level data to estimate hospitalization risks, mortality rate, and COVID-19 transmission. Moreover, several studies (6, 7) indicate racial and ethnic disparity by health systems increases the risk of getting sick, hospitalized, and dying from COVID-19. Future studies should examine vaccination in different races by age group to estimate who should get the vaccine first. Additionally, the data does not include nonpharmaceutical public health control measures, which would be an essential indicator to control daily cases (8).

### Conclusion

The vaccination was started at the beginning of a “3rd wave“in Tennessee, and by December, and January SARS-CoV2 positive cases and hospitalizations increased considerably. This work has concentrated on the Tennessee’s dynamics COVID-19. It concludes that the vaccine should be optimally targeted at the elderly in the first step, indicating that vaccination reduces daily cases for the whole population, while reduces hospitalization and death rate in the older population. This study, consistent with the previous studies (8), shows mRNA Covid-19 vaccines have a protective effect for blocking transmission even after a single dose. This study also indicates that prioritizing the vaccination of the elderly is a practical approach for reducing the number of deaths and hospitalization.

## Data Availability

is publicly available at https://is publicly available at https://www.tn.gov/health/cedep/ncov/data

## Code Availability

The code to perform all analyses described in Methods and Usage Notes sections was written in R-3.6.2 and is publicly available at https://www.tn.gov/health/cedep/ncov/data.

## Author Approval

All authors have seen and approved the manuscript.

## Declaration of Conflicting Interests

The authors declare that there is no conflict of interest.

## Funding/Financial Support

None Reported

## Author Contributions

Ali Roghani designed the study and implemented methods and analyses.

## Notes

### Competing Interest Statement

The authors have declared no competing interest.

### Author Declarations

The data is publicly available.

## Reference

1. Polack F P, Thomas S J, Kitchin N, et al. Safety and efficacy of the BNT162b2 mRNA Covid-19 vaccine N Engl J Med 2020; 383, 2603–15.CrossRefPubMedGoogle Scholar

2. Baden LR, El Sahly HM, Essink B, et al. Efficacy and safety of the mRNA-1273 SARS-CoV-2 vaccine N Engl J Med December 30, 2020. DOI: 10.1056/NEJMoa2035389.CrossRefGoogle Scholar

3. Huang, B., Wang, J., Cai, J., Yao, S., Chan, P. K. S., Tam, T. H. W.,… & Lai, S. (2021). Integrated vaccination and physical distancing interventions to prevent future COVID-19 waves in Chinese cities. Nature Human Behaviour, 1–11.

4. U.S. Census Bureau (2019). American Community Survey 1-year estimates. Retrieved from Census Reporter Profile page for Tennessee http://censusreporter.org/profiles/04000US47-tennessee/

5. Dooling K, Marin M, Megan W, et al. The advisory committee on immunization practices’ updated interim recommendation for allocation of COVID-19 vaccine — United States, December 2020. MMWR 2020; 51:52. https://www.cdc.gov/mmwr

6. Hooper MW, Nápoles AM, Pérez-Stable Ej. COVID-19 and racial/ethnic disparities. JAMA. 2020;323(24):2466–2467.

7. Muñoz-Price Ls, Nattinger AB, Rivera F, Hanson R, Gmehlin CG, Perez A, et al. Racial disparities in incidence and outcomes among patients with COVID-19. JAMA Network Open. 2020;3(9):1–12

8. Moore, S., Hill, E. M., Dyson, L., Tildesley, M., & Keeling, M. J. (2020). Modelling optimal vaccination strategy for SARS-CoV-2 in the UK. medRxiv.

9. Carrillo-Vega, M. F., Salinas-Escudero, G., García-Peña, C., Gutiérrez-Robledo, L. M., & Parra-Rodríguez, L. (2020). Early estimation of the risk factors for hospitalization and mortality by COVID-19 in Mexico. PLoS One, 15(9), e0238905.

